# “How Social Connection and Engagement Relate to Functional Limitations and Depressive Symptoms Outcomes After Stroke”

**DOI:** 10.1101/2023.03.07.23286965

**Authors:** Joanne Elayoubi, William E. Haley, Monica E. Nelson, Gizem Hueluer

## Abstract

**Background:** Stroke commonly leads to disability and depression. Social connection and engagement can be protective against functional decline and depression in the general population. We investigated the effects of social connection and engagement on trajectories of function and depressive symptoms in stroke.

**Methods:** Participants were 898 individuals with incident stroke from the Health and Retirement Study between 1998-2012. Multilevel modeling was used to examine associations of social connection and engagement with changes in functional limitations in instrumental activities of daily living (IADLs) and depressive symptoms over time. Models controlled for age, gender, education, and race/ethnicity. Moderation analyses examined whether high social connection and engagement reduced depressive symptoms for survivors with high IADL impairment.

**Results:** Social connection and engagement were generally associated with fewer IADL limitations and depressive symptoms at the time of stroke and after stroke. For example, participants who felt lonely and did not provide help to others before stroke had more IADL limitations. Pre-stroke volunteering was associated with less increase in IADL limitations with stroke and increase in having friends and providing help to others compared to one’s pre-stroke status were associated with fewer IADL limitations after stroke. For depressive symptoms, participants who felt lonely and did not have a friend or partner before stroke had more depressive symptoms, and participants who had children residing nearby before stroke showed less increase in depressive symptoms. Moderation effects were not found for social connection and engagement on high IADL impairment and depressive symptoms.

**Conclusions:** Findings suggest that social connection and engagement may reduce the negative physical and psychological outcomes of stroke, both at baseline and after stroke. Efforts to enhance social engagement and diminish loneliness may both enhance population well-being and enhance resilience and recovery from stroke and other illnesses.

## Introduction

Stroke is the leading cause of disability in the world^1^ associated with functional limitations and post-stroke depression. The impact of stroke can impede psychological and social functioning for years following stroke^2^. Considerable research has identified factors that may enhance stroke recovery, including speech therapy, strength and aerobic training that improve mobility, balance, gait and cognition^3^.

Social isolation has received increasing attention as a factor associated with heightened depression and poor health outcomes, including increased risk of mortality^4^ in general populations. Studies have also found associations between social isolation and lower gray matter volumes in temporal, frontal, and hippocampal regions of the brain^5^. Social connection, from a *structural* perspective, is the existence of interconnections among different social relationships and roles. Social engagement includes *functions* provided by or perceived to be available because of social relationships^6^.Both can be protective against and enhance recovery from a variety of stressful life circumstances including widowhood^7^. Social connection and engagement have also been shown to be especially valuable after exposures to relatively high levels of stress, known as the buffering effect^8^. However, social connection and engagement have received little emphasis as protective factors that could be advantageous in enhancing stroke outcomes and recovery.

Several studies that examined the role of social connection and engagement in stroke recovery did so using data only after the onset of stroke. A cross-sectional study examining functional status using three-day averages of daily stepping for participants who had experienced a stroke at least six months prior, found social connection indicators, including being married or living with someone, associated with more daily stepping^9^. A prospective study also found social engagement indicators of more frequent social interactions with family and friends immediately at the time of hospital admission were associated with improved activities of daily living (ADLs) performance three months post-stroke^10^. Other studies that examined social engagement found stroke survivors who engaged in work and social activities following stroke had improved function and better perceived recovery during their first year^11^. A longitudinal study found higher levels of social engagement through social support at the time of stroke associated with more improvements in post-stroke depression^12^.

Although these findings concerning social connection and engagement, functional change, and depression in stroke are promising, prior studies have included non-representative samples of stroke survivors recruited from hospital admissions, gathered social connection and engagement data only at post-stroke, and used a narrow range of indicators of social connection and engagement. In a previous publication, we showed that both pre-stroke and post-stroke social connection and engagement predicted better episodic memory after stroke^13^. We know little about how both pre- and post-stroke social connection and engagement may affect post-stroke daily functioning and depressive symptoms, and which dimensions of social connection and engagement (e.g., structural social connection indicators such as marital status, and/or functional social engagement indicators like volunteering) may affect post-stroke outcomes.

Also, more evidence is needed to determine whether social connection and engagement can protect against depressive symptoms resulting from high stroke impairment. The present study addresses these limitations by using data from a representative national sample, the Health and Retirement Study (HRS). Our study also considers a broad range of social indicators of subjective loneliness, structural social connection, and social engagement, measured repeatedly both before and after stroke. We examine longitudinal associations between pre- and post-stroke social connection and engagement on post-stroke changes in daily functioning (instrumental activities of daily living [IADLs]) and depressive symptoms.

We hypothesized higher pre-stroke levels of social connection and engagement, and changes over time indicating higher social connection and engagement after stroke, would be associated with fewer IADL limitations and depressive symptoms at the time of stroke and post-stroke after controlling for relevant covariates. We also hypothesized that higher social connection and engagement would be particularly valuable in buffering against the negative effects of IADL impairment after stroke, such that stroke survivors with relatively high post-stroke IADL impairment would show especially marked benefits of social connection and engagement on reducing depressive symptoms.

## Methods

### Procedure and Participants

Data were taken from the HRS, a nationally representative population-based study of households in the United States that include at least one member who is 50 years old or older. Spouses/partners who live in the same household were invited to participate regardless of age. HRS includes data from 40,000+ individuals. Data are collected every two years. Detailed information about the procedure and participants were previously provided^14^. Information relevant to the present study is reported below.

To study the associations between social connection and engagement and trajectories of functional limitations and depressive symptoms surrounding stroke, we included all participants who reported incident stroke in their HRS interviews between 1998-2012.We used data from 1998 onwards because that was the year older cohorts joined HRS, and we did not include data from years 2014 and 2016 because specific social variables, including having friends or relatives, and frequency of social visits were measured differently. We also excluded participants who resided in a nursing home. We used data from participants who provided the exact date of stroke (month and year) and had at least two data points, one taken pre-stroke and one taken post-stroke. We identified 898 participants who met these criteria, and they contributed 5,058 longitudinal observations. On average, participants contributed 5.6 longitudinal observations (*SD*=1.8, range=2 to 8).

### Measurements

#### Incident Stroke

The timing of incident stroke was determined by self-report of a first stroke and not by medical reports or hospitalization records. Previous studies have shown that self-reported stroke is an accurate measure for stroke incidence and compatible with hospital records^15^. Participants self-reported at each wave whether they ever had a stroke. In addition, participants provided the date of stroke (month/year), which was used to calculate two-time metrics. The first-time metric, time to/from stroke ranged from -12.9 to 14.1 years and was used to describe trajectories of functional limitations and depressive symptoms as a function of the distance from the time of stroke, which was coded as 0 and served as a baseline. As a result, years prior to stroke had negative values, indicating time-to-stroke and years after stroke had positive values, indicating time-from-stroke. The second time metric, post-stroke, was used to examine differences in pre-versus post-stroke time periods. All observations before incident stroke were coded as 0 and all observations after incident stroke were coded as 1.

#### Functional Limitations

Functional limitations were assessed as difficulties with IADLs. Participants first indicated whether or not they had difficulty on any of the five IADLs (“yes”=1; “no”=0) including using a telephone, taking medication, handling money, shopping, and preparing food (range: 0 to 5) as done in prior research using the same dataset^16^.

#### Depressive Symptoms

Depressive symptoms were assessed with seven items from the 8-item Center of Epidemiologic Studies Depression (CES-D) scale^17^ across all waves. One item from the scale (loneliness) was dropped from the sum score because loneliness was used to indicate social engagement, which has been done in research on loneliness^18^.

#### Social Connection

Following widely cited conceptual approaches in this field^6^, social connection was represented by using five structural indicators including household size, having friends and relatives in one’s neighborhood, partnered status, and presence of children residing within 10 miles. Household size indicated the number of individuals living in the same household (range: 1 to 11). Having friends and relatives were each binary coded (“yes”=1; “no”=0). Partnered status was coded as 1 if participants were married, married with spouse absent, or partnered, and 0 if participants were separated, divorced, widowed, or never married. Children residing within 10 miles was binary coded (“yes”=1; “no”=0) based on whether participants had (a) any children, (b) co-resident children, and, (c) children living within 10 miles.

#### Social Engagement

Social engagement was represented by four functional indicators: loneliness, volunteering in the community, frequency of social visits to friends or relatives in the neighborhood and providing unpaid help to friends and/or relatives in the neighborhood. For loneliness, participants were asked if they felt lonely in the previous week (“yes”=1; “no”=0).

Volunteering in the past 12 months and providing unpaid help were binary coded (“yes”=1; “no”=0), and frequency of social visits was coded as daily or more frequent=5, at least weekly=4, at least every two weeks=3, at least monthly=2, at least yearly=1, almost never=0.

#### Covariates

Covariates included age (in years), years of education (range=0-17+ years), gender (male=1 and female=2), and race/ethnicity. Racial/ethnic categories included non-Hispanic White=1(76%); non-Hispanic Black=2 (15%) and Other=0 (5.01% Hispanic White, 0% Hispanic Black, 2.00% non-Hispanic Other, 2.00% Hispanic Other).

### Statistical Analyses

Trajectories of IADL limitations and depressive symptoms surrounding stroke were examined with multilevel models^19^. This type of analysis allows for examination of both within-person change (i.e., changes found in individuals over time) and between-person differences (i.e., differences between people at baseline and over time). Within-person change can occur both due to aging and due to events like stroke. Analysis of trajectories allows examination of both differences in baseline levels and the rate of change in functional limitations and depressive symptoms for different individuals. First, we estimated models that only included the effects of time in relation to the stroke event and no other predictors on functional limitations and depressive symptoms. The main analyses of interest examined the effects of pre-stroke levels and within-person changes from pre-stroke levels of (a) social connection and (b) social engagement variables on functional limitations and depressive symptoms. Within-person change was calculated as how much each person varies at each time point compared to their mean level of pre-stroke social connection and engagement^20^. These analyses also controlled for age, gender, education, and race/ethnicity centered at sample means. Additional details on methodology are available in Supplemental Materials. Moderation analyses assessed whether the effects of social connection and engagement variables on post-stroke depressive symptoms differed as a function of IADL limitations. We included interaction terms between the pre-stroke levels of social connection and engagement variables and the within-person variables indicating changes in IADL limitations from pre-stroke. All analyses were conducted with PROC Mixed^21^ in SAS software^22^ Version 9.4 with incomplete data treated as missing at random^23^ and statistical significance assessed at *p*<.05 (2-sided).

## Results

### Sample Characteristics

Table 1 shows descriptive statistics for all study variables taken at the earliest observation after the stroke event. Mean age was 72 years (*SD*=10); 46% were male; and 76% were non-Hispanic White. Participants reported 12 years of education on average (*SD*=3.08).

**Table 1.** Baseline Descriptive Characteristics of Study Variables.

| Variables | *M or N* | *SD or %* |
| --- | --- | --- |
| Age at Stroke (Years) | 72.43 | 10.26 |
| Gender |  |  |
| Male | 415 | 46% |
| Female | 483 | 53% |
| Race/Ethnicity |  |  |
| Non-Hispanic White | 683 | 76% |
| Non-Hispanic Black | 135 | 15% |
| Other (Hispanic White, non-Hispanic Other, Hispanic Other) | 80 | 9% |
| Education (Years) | 12.18 | 3.08 |
| Volunteering in the Community |  |  |
| No | 696 | 77% |
| Yes | 202 | 22% |
| Loneliness<sup>a</sup> |  |  |
| No | 654 | 72% |
| Yes | 244 | 27% |
| Household Size | 2.09 | 1.11 |
| Children Residing Within 10 Miles |  |  |
| No | 317 | 35% |
| Yes | 581 | 64% |
| Partnered Status |  |  |
| Not Married or Cohabiting | 387 | 43% |
| Married or Cohabiting | 511 | 56% |
| Friends Living in Neighborhood |  |  |
| No | 305 | 33% |
| Yes | 593 | 66% |
| Relatives Living in Neighborhood |  |  |
| No | 604 | 67% |
| Yes | 294 | 32% |
| Providing Unpaid Help to Friends or Relatives in Neighborhood |  |  |
| No | 576 | 64% |
| Yes | 322 | 35% |
| Social Visits with Friends or Relatives in Neighborhood<sup>b</sup> | 2.48 | 1.97 |
| Depressive Symptoms<sup>c</sup> | 2.00 | 2.10 |
| Instrumental Activities of Daily Living<sup>d</sup> | 0.83 | 1.35 |

*Note.* $n = 898$ . Abbreviations: M = mean. SD = standard deviation. <sup>a</sup>Measured as the perceived loneliness item from the 8-item Center for Epidemiologic Studies Depression (CES-D) scale.
<sup>b</sup>Frequency of social visits with friends or relatives in the neighborhood (0 = almost never, 1 = at least yearly, 2 = at least monthly, 3 = at least every two weeks, 4 = at least weekly, 5 = daily or more frequent). <sup>c</sup>Measured with the 8-item CES-D scale with the perceived loneliness item removed. <sup>d</sup>Measured as a sum of difficulty (0 = no, 1 = yes) with using a telephone, taking medication, handling money, shopping, and preparing meals (range: 0-5).

### Trajectories of Functional Limitations and Depressive Symptoms Surrounding the Stroke Event

First, we examined the average trajectories of functional limitations and depressive symptoms surrounding the stroke event. In Figure 1A for IADL limitations, participants on average showed increases over time (Supplemental Table 1). Figure 1B illustrates that for depressive symptoms, participants showed small increases over time before stroke, a substantial increase at the time of stroke, and a decline over time in the post-stroke period (Supplemental Table 2).

**Figure 1:**
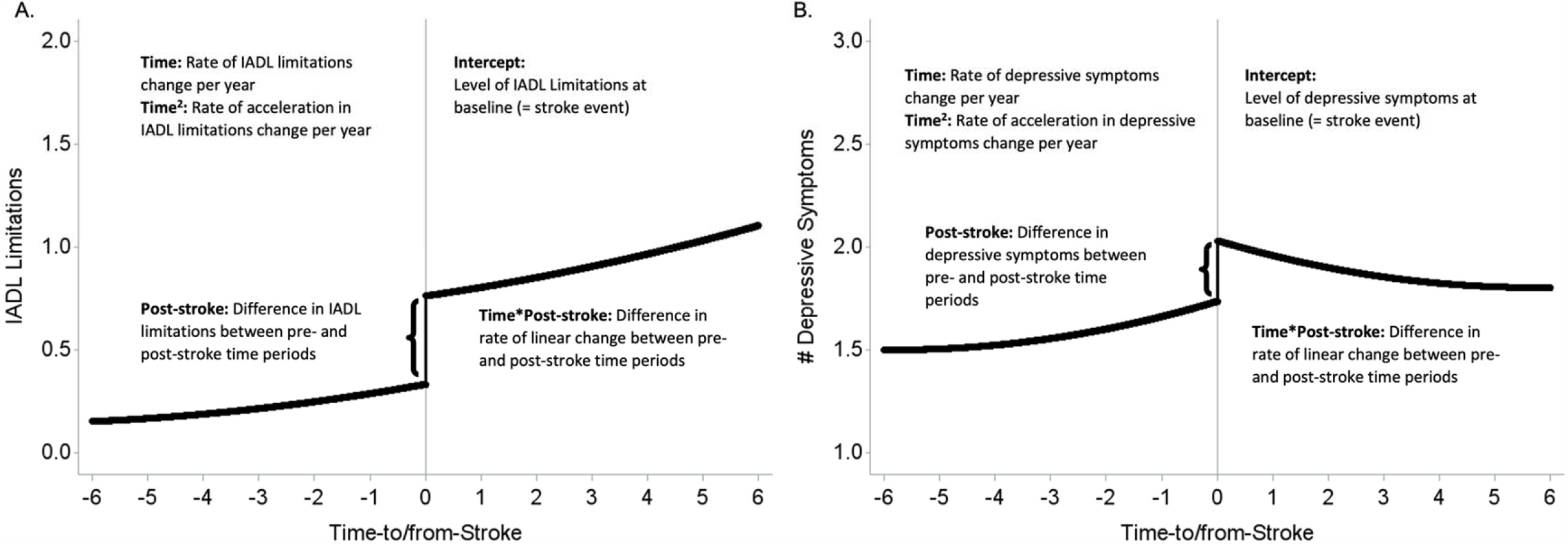
Average trajectory of IADL limitations and depressive symptoms surrounding the incident stroke event. *Note*. A) Trajectory of IADL limitations surrounding stroke (6years= average number of longitudinal observations per participant). Participants had more limitations in IADLs in the post-stroke period compared with the pre-stroke period. There was an accelerated rate of change per year in IADL limitations. B) Trajectory of depressive symptoms surrounding stroke. Participants had more depressive symptoms in the post-stroke period compared with the pre-stroke period.

### The Relationship of Social Connection and Engagement to Changes in Functional Limitations with Incident Stroke

Table 2 shows the results from analyses examining the role of social connection and engagement variables on IADL limitations while controlling for age, education, race/ethnicity, and gender. In analyses examining social connection variables, there was a significant interaction between within-person change in friendship and post-stroke, such that having friends compared to pre-stroke friendship status was associated with significantly fewer limitations in IADLs in the post-stroke period (β=-0.13, SE=0.06, *p*=.0231). This interaction effect indicated that the effect of friendship becomes more important for IADL limitations after stroke compared to pre-stroke. For household size, participants with larger household sizes in the pre-stroke period had significantly more post-stroke increase in IADL limitations (γ=0.11, SE=0.05, *p*=.0248), but compared to smaller household sizes, there were no differences in the rate of change in the post-stroke period (see Figure 2A).

**Table 2.** Results from Multilevel Models Examining Effects of (a) Social Connection and (b) Social Engagement on IADLs.

| Predictors | Pre-Stroke<br>Average of<br>Variable<br>Predicting<br>Baseline<br>IADLs |  | Pre-Stroke<br>Average of<br>Variable<br>Predicting Linear<br>IADL Change<br>over Time |  | Pre-Stroke<br>Average of<br>Variable<br>Predicting<br>Change with<br>Stroke in<br>IADLs |  | Pre-Stroke<br>Average of<br>Variable<br>Predicting Post-Stroke Change<br>over Time in<br>IADLs |  | Within-Person<br>Change in<br>Variable from<br>Pre-Stroke<br>Average<br>Predicting<br>IADLs |  | Within-Person<br>Change in<br>Variable from<br>Pre-Stroke<br>Average<br>Predicting<br>IADLs in Post-Stroke Period |  |
| --- | --- | --- | --- | --- | --- | --- | --- | --- | --- | --- | --- | --- |
|  | Estimate | SE | Estimate | SE | Estimate | SE | Estimate | SE | Estimate | SE | Estimate | SE |
| *Social Connection* |  |  |  |  |  |  |  |  |  |  |  |  |
| Household Size | 0.06 | 0.03 | -0.01 | 0.01 | **0.11** | 0.05 | 0.00 | 0.01 | 0.01 | 0.03 | 0.03 | 0.04 |
| Children Residing Within 10 Miles | -0.08 | 0.07 | 0.00 | 0.01 | -0.01 | 0.11 | 0.00 | 0.02 | -0.01 | 0.06 | -0.02 | 0.08 |
| Partnered Status | -0.04 | 0.07 | 0.00 | 0.01 | -0.09 | 0.11 | -0.01 | 0.02 | -0.01 | 0.08 | 0.07 | 0.11 |
| Friends Living in Neighborhood | -0.06 | 0.08 | -0.01 | 0.02 | 0.08 | 0.12 | 0.01 | 0.02 | -0.03 | 0.04 | **-0.13** | 0.06 |
| Relatives Living in Neighborhood | 0.07 | 0.07 | 0.00 | 0.01 | 0.07 | 0.12 | -0.01 | 0.02 | 0.05 | 0.05 | 0.01 | 0.06 |
| *Social Engagement* |  |  |  |  |  |  |  |  |  |  |  |  |
| Loneliness | **0.43** | 0.09 | 0.02 | 0.02 | 0.03 | 0.14 | -0.01 | 0.02 | 0.07 | 0.05 | 0.09 | 0.07 |
| Providing Unpaid Help to Friends<br>or Relatives in Neighborhood | **-0.23** | 0.08 | -0.01 | 0.02 | -0.10 | 0.13 | -0.02 | 0.02 | 0.00 | 0.04 | **-0.19** | 0.06 |
| Social Visits to Friends or Relatives<br>in Neighborhood | -0.02 | 0.02 | 0.00 | <0.01 | -0.01 | 0.03 | 0.01 | <0.01 | -0.01 | 0.01 | -0.01 | 0.01 |
| Volunteering in Community | 0.00 | 0.07 | -0.01 | 0.01 | **-0.22** | 0.12 | **0.05** | 0.01 | -0.06 | 0.05 | -0.07 | 0.07 |

*Note.* $n = 898$ . IADLs = Instrumental activities of daily living. SE = Standard error. Analyses controlled for age at time of stroke, gender, race/ethnicity, and years of education. Pre-stroke averages were centered at the sample mean. Within-person change variables indicate deviations from the pre-stroke average at each time point. Bolded values indicate significant results at $p < .05$ .

**Figure 2:**
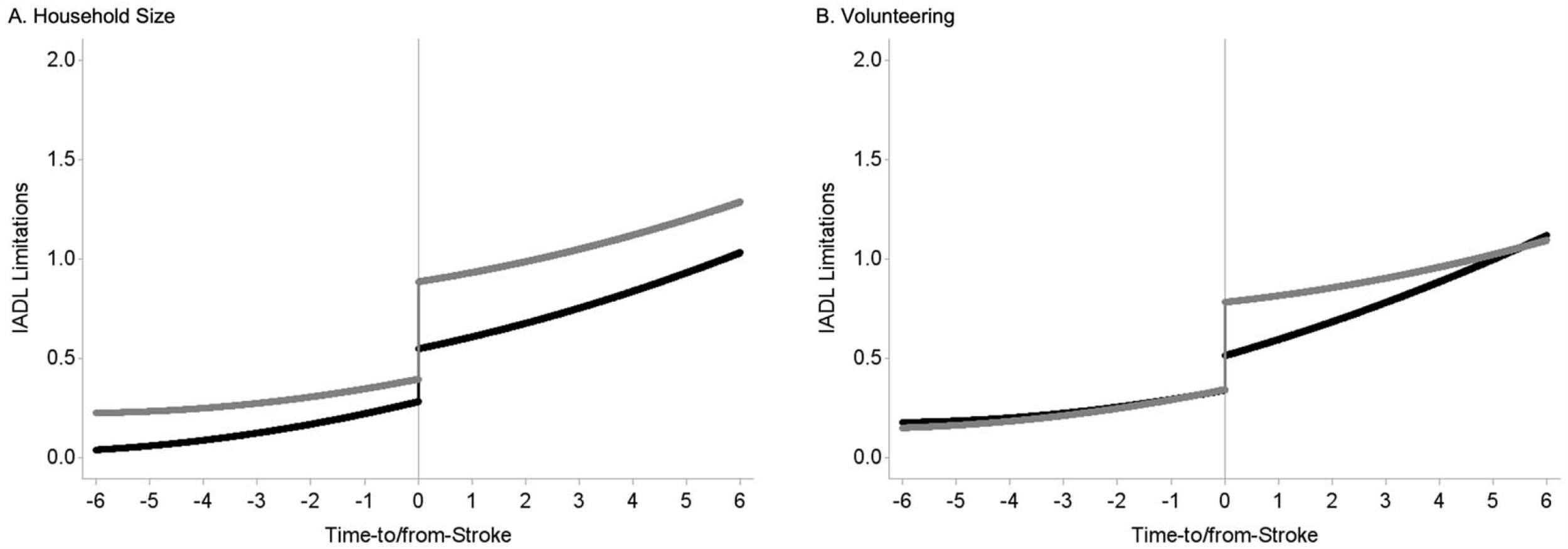
Social connection and engagement variables and IADL limitations surrounding stroke. *Note*. A) Compares trajectory of IADL limitations surrounding stroke (6years= average number of longitudinal observations per participant) for individuals with small household size =1 (black line) versus large household size=3 (gray line). Participants with larger household sizes at pre-stroke had significantly more post-stroke increase in IADL limitations. There were no differences in the rate of change in IADL limitations in the post-stroke period between large and small household sizes. B) Compares trajectory of IADL limitations surrounding stroke for participants who volunteered at pre-stroke (black line) versus participants who did not volunteer at pre-stroke (gray line). Participants who volunteered during the pre-stroke period had less increase in IADL limitations with stroke, but more increase in IADL limitations over time.

In analyses examining social engagement variables, perceived loneliness and providing help were significantly associated with IADL limitations at baseline, with participants who were lonely before stroke having significantly more IADL limitations (γ=0.43, SE=0.09, *p*<0.001), and participants who provided help before stroke having significantly fewer IADL limitations at baseline (γ=-0.23, SE=0.08, *p*=.0043). Also, providing help was associated with fewer IADL limitations after stroke (β=-0.19, SE=0.06, *p*=.0015). Participants who volunteered before stroke had less increase in IADL limitations with stroke (γ=-0.22, SE=0.12, *p*=.0269), but more increase in IADL limitations over time (γ=0.05, SE=0.01, *p*=.0065) (see Figure 2B).

### The Relationship of Social Connection and Engagement to Changes in Depressive Symptoms with Incident Stroke

Table 3 shows the results from analyses examining the role of social connection and engagement variables on depressive symptoms while controlling for age, education, race/ethnicity, and gender. In analyses examining the role of social connection variables, participants who were partnered (γ=-0.41, SE=0.15, *p*=.0414) and had friends (γ=-0.34, SE=0.16, *p*=.0372) had significantly fewer depressive symptoms at baseline. Although participants with children residing within 10 miles did not have significantly fewer depressive symptoms at baseline, they did have less of an increase in depressive symptoms over time compared to participants without children residing within 10 miles (γ=-0.05, SE=0.02, *p*=.0300) (see Figure 3). Moreover, becoming partnered was associated with fewer depressive symptoms (β=-0.41, SE=0.19, *p*=0.023).

**Table 3.**
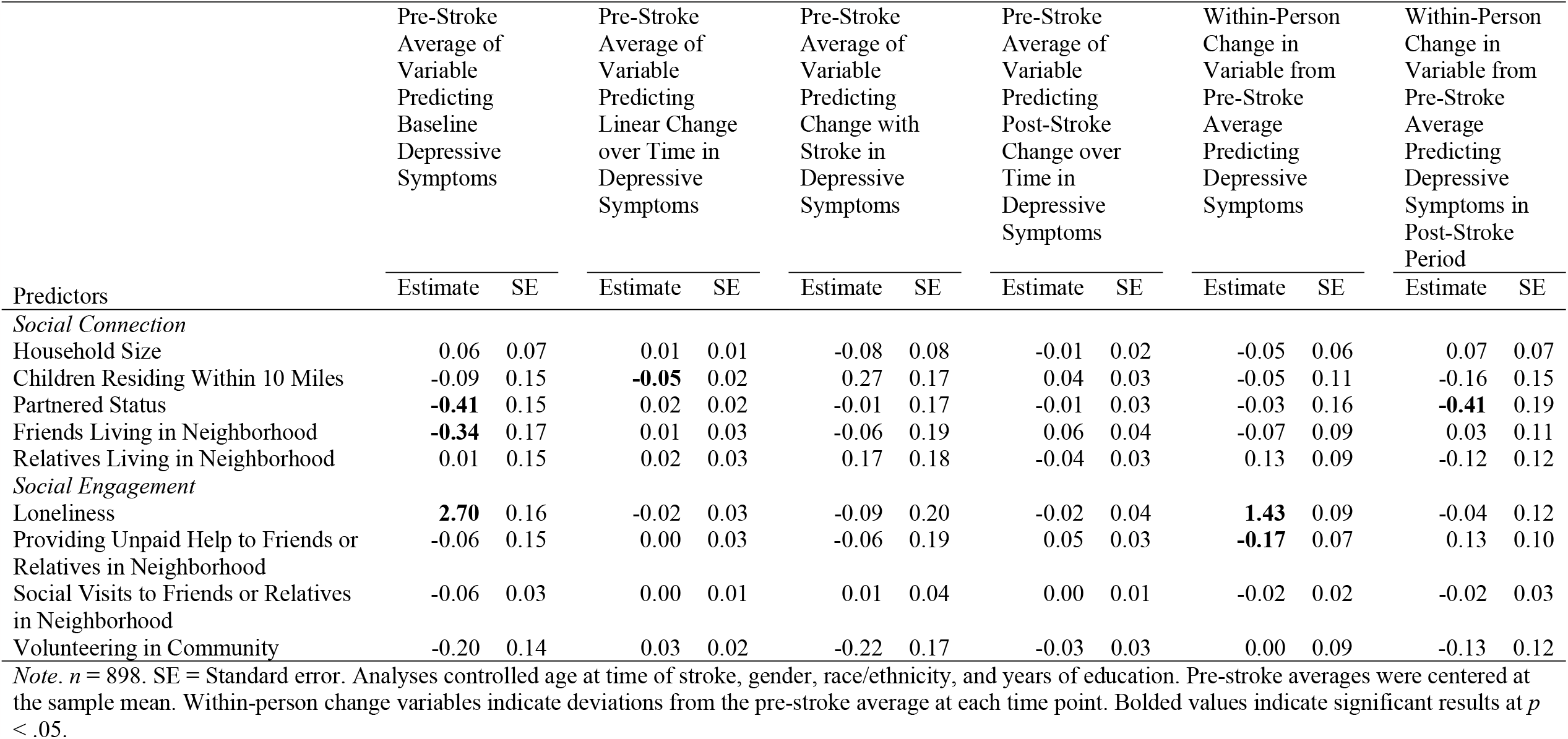
Results from Multilevel Models Examining Effects of (a) Social Connection and (b)Social Engagement on Depressive Symptoms.

**Figure 3:**
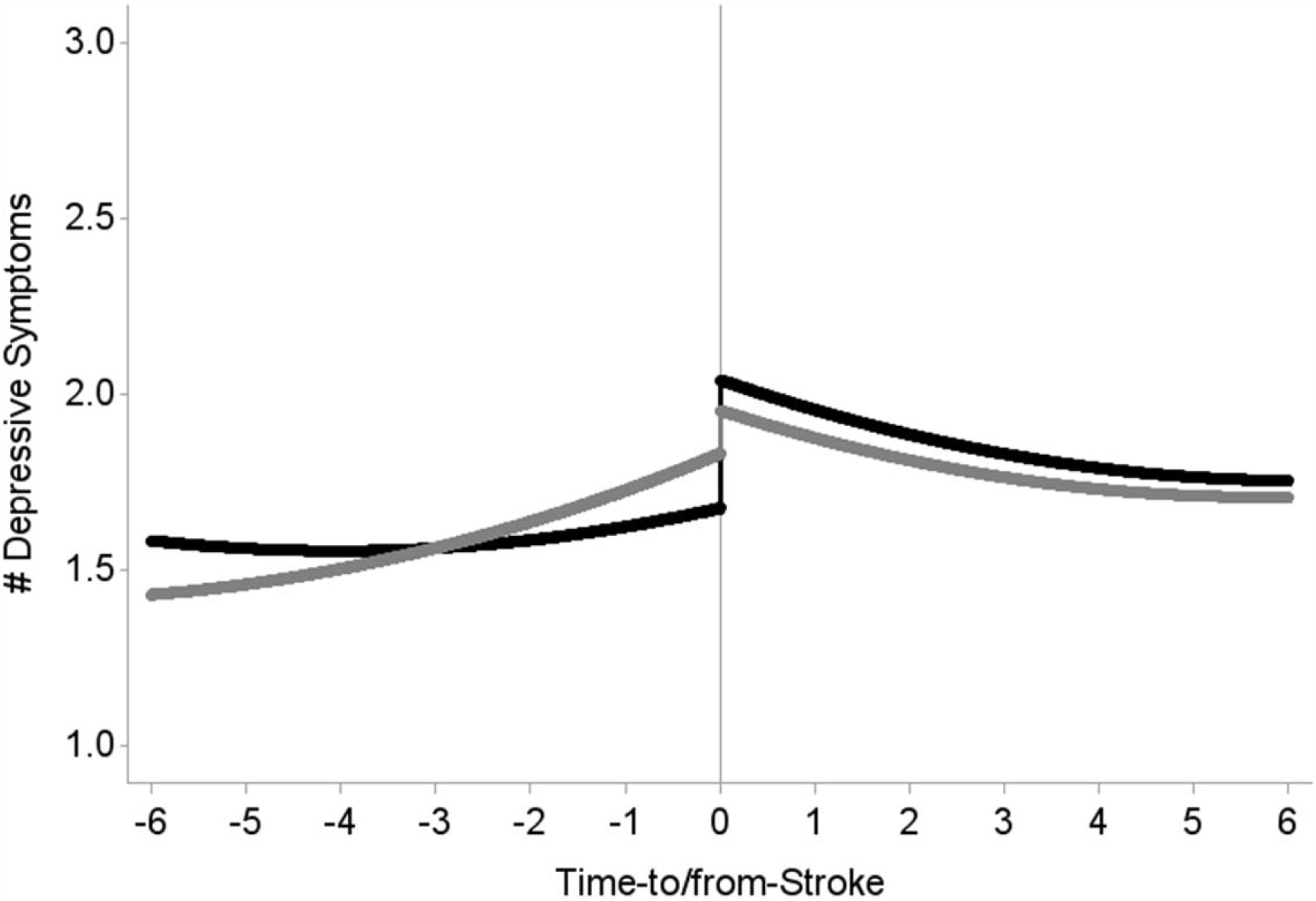
Depressive symptoms surrounding stroke for participants with and without children residing nearby *Note*. Compares trajectory of depressive symptoms surrounding stroke (6years= average number of longitudinal observations participants contributed to the study) for participants with children residing nearby (black line) and without children residing nearby (gray line). Participants with children residing within 10 miles did not have significantly fewer depressive symptoms at baseline but had less of an increase in depressive symptoms over time compared to participants without children residing within 10 miles.

In analyses examining social engagement variables, pre-stroke loneliness was significantly associated with more depressive symptoms at baseline (γ=2.70, SE=0.16, *p*<.0001). and feeling lonelier was associated with significantly more depressive symptoms at a given time (γ =1.43, SE=0.09, *p*<.0001). Increases in providing help were associated with fewer depressive symptoms at a given time point (γ =-0.17, SE=0.07, *p*=.0350).

### Moderations of the Functional Limitations-Depressive Symptoms Relationship by Social Connection

We did not find any moderation effects of social connection and engagement on change in functional limitations and depressive symptoms after covariate adjustment (see Supplemental Table 3).

## Discussion

There is extensive literature documenting the importance of social connection and engagement on psychological well-being and lower mortality^18^, improving functional ability and reducing depression^24,18^. However, we have not identified any previous studies that examined an extensive range of pre- and post-stroke measurements of social connection and engagement (approximately 13 years prior and 14 years after) and their association with post-stroke impairment and depressive symptoms. This study does so and provides evidence that social connection and social engagement may protect against increases in functional limitations and depressive symptoms with incident stroke.

In line with our hypothesis, we found significantly fewer IADL limitations among participants who provided help and did not feel lonely before stroke. We also found better post-stroke trajectories in IADLs for participants who developed friendships and provided help in the post-stroke period. Our results suggest that these structural social connection and functional social engagement factors may play a crucial role in providing resilience to post-stroke impairment.

With respect to stroke survivors’ trajectories of depressive symptoms, in support of our hypothesis, participants who at pre-stroke were not partnered, felt lonely, or did not have friends in their neighborhood reported more depressive symptoms at the time of stroke. In addition, participants who had children residing within 10 miles before stroke occurrence reported less increase in depressive symptoms over time. Moreover, participants who became less lonely and provided more help reported fewer depressive symptoms, and participants who became partnered reported fewer depressive symptoms post-stroke.

Taken together, our findings show that both strong structural measures of social connection and social engagement including perceived loneliness and other all have value in protecting against both functional decline and depressive symptoms among older adults with stroke. This study highlights the need for future studies to examine whether building social connections with friends, children, and partners and engaging in helping behaviors before and after stroke can reduce functional decline and depressive symptoms. Despite these positive findings, participants with larger households had more increase in IADL with stroke. A possible reason may be that those who were able to live independently before stroke had better health and functioning and health differences may not have been fully captured using this variable. Another possible explanation for this finding may be that participants with large household sizes have more people surrounding them to perform these activities after their stroke compared to participants with small household sizes. Also, participants who volunteered before stroke had less increase in IADL with stroke but more increase post-stroke. It may be that volunteering is protective of functional health with stroke but the maintenance of these benefits over time becomes more difficult as participants deal with post-stroke changes.

Intervention studies have examined related issues and provided results consistent with our findings. Interventions using home-based telerehabilitation for stroke found older adults with larger social networks during treatment had better motor function after twelve weeks^25^ compared to a historical cohort control of stroke patients who did not receive telerehabilitation. The study suggests that treatment can be more effective when there are others present to reinforce and support rehabilitation. Also, a physical activity intervention for older adults with major disabilities due to a variety of conditions^26^ found older adults who had high social participation at baseline, in terms of visiting family and friends or attending group events, had better motor function 42 months post-intervention compared to older adults randomized into a health education program. Older adults with limited social participation did not improve in mobility from the physical activity intervention compared to the health education group. The study suggests baseline social participation as a significant factor in rehabilitation for delaying mobility disability.

A randomized controlled trial examining the effects of a social support intervention on post-stroke depression found that stroke patients who received routine rehabilitation that was supplemented by a social support intervention showed significantly reduced depressive symptoms eight weeks post-stroke^27^ compared to a control group of stroke patients that received routine rehabilitation only. This prior study shows social support as a vital factor in keeping stroke patients motivated and adherent to the rehabilitation process.

Our study provided valuable new information by using pre- and post-stroke measures of social connection and engagement and examining their association with IADLs and depressive symptoms over time. Thus, we were able to examine changes and distinguish between changes likely due to aging alone, plus the added impact of stroke. However, our study had several limitations. We did not examine these associations over recurrent or multiple strokes. Stroke was self-reported, and we did not have information on different types of strokes (e.g., hemorrhagic vs. ischemic), stroke fatalities, stroke severity, or a modified Rankin score to measure post-stroke disability. Similar studies supported by medical records may provide a clearer understanding of functional limitations and depressive symptoms in stroke survivors. The sample lacked racial and ethnic diversity, a special concern since there are well-known racial and ethnic differences in stroke and stroke outcomes^28^. Finally, IADL limitations and depressive symptoms were measured every two years in HRS. More closely spaced observations of IADL limitations and depressive symptoms would be useful in examining the immediate effects of stroke more accurately.

Despite these shortcomings, our study used a broad context of social indicators to frame social connection and engagement, pre-stroke data, and a high number of observations per participant. Social connection and engagement interventions have been demonstrated to be effective at enhancing social connection and leading to related benefits in well-being. The development of a “Connection Plan” for older patients^29^ and training techniques in accessing social networking websites^30^ are both promising programs and relevant for building social ties. A recent randomized trial documented that a simple intervention using phone calls to isolated older adults during COVID19 helped reduce anxiety, depression, and loneliness^31^. Such interventions that have been used in broader populations of older adults could be adapted and may prove especially beneficial for older adults who have had a stroke and are socially vulnerable (e.g., lonely, no friends or partners).

This study is also relevant to assessment, discharge planning, and rehabilitation for stroke survivors. Discharge planning and stroke rehabilitation commonly include involvement of family caregivers^32^, but a thorough assessment of stroke survivors’ social connection and engagement that include plans for reengaging with others could be valuable in treatment planning. Enhancing social connection and engagement as a part of stroke rehabilitation can also make important contributions for people with stroke. For instance, a pilot study investigating the effects of group singing in a community choir enhanced mood, global functioning, cognition, and sense of belonging for people with aphasia at a 20-week follow-up^33^. Also, a recent set of best practice recommendations suggested broader community participation as a means of reintegrating people with stroke into their communities^3^ in addition to rehabilitation and family involvement.

In conclusion, findings from this study highlight the importance for future researchers to examine whether modifications towards greater social connection and engagement can decrease functional impairment and depressive symptoms with stroke. Post-stroke social engagement beyond rehabilitation sessions could prove valuable for community reintegration.

## Data Availability

The study is based on publicly available data and interested investigators can access the data via the HRS website: https://hrsonline.isr.umich.edu.

## Acknowledgements

We would like to thank Dr. Mohamad S. Saad for his general advice and guidance and Dr. Debra Dobbs for her help in editing the first draft of the manuscript.

## Disclosures

Joanne Elayoubi, William E. Haley, Monica E. Nelson, and Gizem Hueluer report no disclosures.

Supplemental Material: Section 1, Table 1, Table 2, and Table 3

